# *PTPN1*-related autoinflammation is a common cause of Aicardi-Goutières Syndrome with reduced penetrance

**DOI:** 10.64898/2026.03.27.26345228

**Authors:** Daniel G. Calame, Emma K. Wiener, Francesco Gavazzi, Anjana Sevagamoorthy, Amy Pizzino, Kaley Arnold, Carlos Dominguez Gonzalez, Tejas Jammihal, Mariko Bennett, Laura Adang, Sarah Woidill, Matthew T. Whitehead, Arastoo Vossough, Russell D’Aiello, Asako Takanohashi, Janhavi Lele, Cas Simons, Rocio Rius, Edward Formaini, Kathleen E. Sullivan, Milena Andzelm, Darius Ebrahimi-Fakhari, Catherine Otten, Stephen Wong, Thomas Reynolds, Raphael Schiffmann, Nicole I. Wolf, Quinten Waisfisz, Jikke-Mien Niermeijer, Danielle DeMarzo, Moez Dawood, Mira Gandhi, Jesse M. Levine, Ivan K. Chinn, Kristen Fisher, Lisa Emrick, Chadi Al Alam, Rauan Kaiyrzhanov, Reza Maroofian, Henry Houlden, Shalini N. Jhangiani, Heer H. Mehta, Donna M. Muzny, Fritz J. Sedlazeck, Jennifer E. Posey, James R. Lupski, Richard A. Gibbs, Ramakrishnan Rajagopalan, Adeline Vanderver

## Abstract

**Purpose:** Aicardi-Goutières syndrome (AGS) is a type I interferonopathy presently associated with nine genes. *PTPN1* is a negative regulator of the interferon pathway previously associated with chronic inflammation and recently type 1 IFN autoinflammation.

**Methods:** Genomic data from undiagnosed individuals with suspected AGS were interrogated for *PTPN1* variants, and predicted loss-of-function (pLOF) and damaging missense variants in *PTPN1* were sought in two additional academic databases as well as the *All of Us* database.

**Results:** We identified 13 cases with ultra-rare heterozygous pLOF or highly damaging missense variants in *PTPN1*. Nine cases were identified in a cohort of 53 individuals (∼ 17%) with clinical, imaging and persistent biochemical features of AGS. Median age of onset is 1.75 years (IQR 0.67), significantly later (p< 0.0001) than other AGS genotypes. Four additional cases were identified in academic datasets with variable clinical features suggestive of autoinflammation. Additionally, 49 individuals with ultra-rare, damaging *PTPN1* variants were identified in the *All of Us* database, none had features suggestive of AGS, but autoimmunity was highly prevalent (∼21.6%).

**Conclusion:** Our data implicate *PTPN1* as a cause of later-onset presentations of AGS within a broader spectrum of autoinflammatory phenotypes. Segregation and biobank data demonstrate reduced penetrance, with carriers being enriched for autoimmune disorders.

## Introduction

Aicardi-Goutières syndrome (AGS) is a treatable type I interferonopathy resulting from dysregulation of anti-viral type I interferon signaling pathways. Nine genes to date have been definitively associated with AGS due to aberrant upregulation of the type I interferon response triggered by self-DNA/RNA: *TREX1*, *RNASEH2A*, *RNASEH2B*, *RNASEH2C*, *SAMHD1, ADAR*, *IFIH1, RNU7-1* and *LSM11*.^1–7^ Additionally, variants in two genes encoding negative regulators of type 1 interferon signaling, *STAT2* and *USP18*, have been implicated in some cases of severe AGS-like presentations. Such negative regulators are necessary to maintain the balance between adequate antiviral responses and autoinflammation.

Protein tyrosine kinases like the Janus kinases (JAK1, 2, 3 and TYK2) are integral components of the type 1 interferon signaling pathway **(Supplemental Figure 1**). Protein Tyrosine Phosphatases (PTPs) are thus an important check on kinase phosphorylation downstream of interferon receptor activation and negatively regulate the interferon signaling pathway. ^9^ One such PTP, PTP1B, is a cytoplasmic non-transmembrane type PTP encoded by *PTPN1*. PTP1B is a key regulator of innate immunity through its regulation of type I interferon signaling. It has been identified in most innate immune cells, including microglia, and has well-recognized functional roles both in homeostasis and disease. ^10^ Dysregulation of PTP1B has been linked to chronic inflammatory pathologies such as asthma, colitis, and cardiovascular disease. ^11–14^ PTP1B also has an established association with Type 2 diabetes due to its interferon-independent regulation of insulin and leptin signaling as well as immune cell activation driving the inflammation of the metabolic syndrome. ^15^

Recently, ultra-rare *PTPN1* missense or predicted loss-of-function (pLOF) variants were identified as a cause of pediatric autoinflammatory encephalopathy in twelve cases, associated with enhanced type I IFN signaling in blood and cerebrospinal fluid (CSF). Importantly, the *PTPN1* variants occurred *de novo* in only three individuals, whereas in nine individuals they were inherited from an unaffected parent. These data combined with *PTPN1*’s pLOF constraint in gnomAD v4.1.1 (pLI=1, LOEUF=0.32) thus suggest *PTPN1* haploinsufficiency causes a type 1 interferonopathy with reduced penetrance, although replication is clearly needed to substantiate the disease association.

Given the emerging evidence that loss-of-function variants in *PTPN1* increase activation of the type I interferon pathway, we investigated a large cohort of patients suspected of having AGS but without a molecular diagnosis for *PTPN1* variants. We find that *PTPN1* variants likely explain a substantial subset of molecularly unresolved AGS cases. We further characterize the clinical features of *PTPN1*-related AGS including response to the JAK inhibitor baricitinib. Finally, we integrate variant segregation data from our cohort, published cases^16^ and the *All of Us* biobank to estimate the penetrance of *PTPN1*-related autoinflammation.

## Methods

### Patient enrollment across cohorts

Affected individuals were recruited from four sources: The Children’s Hospital of Philadelphia (CHOP) as part of the Global Leukodystrophy Initiative Clinical Trial Network (GLIA-CTN) (IRB #14–011236), the Baylor College of Medicine-Genomics Research to Elucidate the Genetics of Rare diseases (BCM-GREGoR) research center (IRB #H-29697), the Amsterdam Leukodystrophy Center, Amsterdam University Medical Center (AUMC)(IRB #2018.300) and the University College London Queens Square Institute of Neurology (UCL IoN) neurogenetics initiative (IRB #07/Q0512/26). All individuals provided written informed consent for publication of clinical and genetic data. For individuals (N=9, LD_0474/ BH15159-1, LD_1504, LD_1289, LD_1433, LD_2020, LD_2444, LD_2578/ BH16506-1, LD_2933, LD_3081) enrolled in the Myelin Disorder Biorepository Project (MDBP) at CHOP, clinical records and clinically ordered or research-generated genome and exome sequences were stored for reanalysis in the MDBP’s data system. The nine MDBP cases were additionally part of a sub-cohort in MDBP of 53 individuals who had a clinical diagnosis of AGS but without a definitive molecular diagnosis after review for other known causes of primary interferonopathy. Two of the MDBP cases were also enrolled in the BCM-GREGoR. Additional individuals were enrolled at the Amsterdam Leukodystrophy Center at the Amsterdam University Medical Center (UMC) (n=2, AGS-41 and AGS-43) or the UCL IoN (n=2, SYNS-04513 and SYNS-05431). Study data were collected and managed using REDCap® electronic data capture tools hosted at The Children’s Hospital of Philadelphia.^17,18^ All patient IDs are not known to anyone outside the research group.

### Clinical Data Extraction across cohorts

The MDBP *PTPN1-*related AGS cohort underwent deep clinical phenotyping using medical records as source documentation. Data on demographics, genotype, age at disease onset, clinical manifestations, and chronological age at the last evaluation were derived from records. Key medical events, including age at presentation and involvement of systemic complications, were extracted using a standardized approach and word bank. For each clinical finding, an age of onset was calculated, or a date approximation was used depending on data availability (for example, if only month or season was mentioned, the first day of the relevant period was considered). A symptom or event was considered noted if it was mentioned in their medical records. A symptom or event was considered not noted if a condition was explicitly excluded based on documentation in the medical record. A symptom or event was considered unknown if not mentioned in the medical record. The timing and dose of baricitinib clinical treatment, when used, was noted. Evidence of supportive clinical laboratory testing, such as CSF neopterin/tetrahydrobiopterin or monocyte expression of interferon stimulated epitopes, was noted where performed.

Review of brain magnetic resonance imaging (MRI) was performed by two pediatric neuroradiologists in consensus. The following parameters were assessed in all cases: Presence of calcifications, cerebral atrophy, deep gray matter atrophy, cerebellar atrophy, any reversible atrophy, brain signal abnormalities, contrast enhancement, and presence of temporal or germinolytic cysts.

Clinical data from non-MDBP cases was collected as available from collaborating clinical geneticists and/or neurologists using a standardized template. Brain MRIs were collected when possible and were reviewed by neuroradiologists as described above. Precise ages were removed from manuscript per medRxiv requirements but are available from the corresponding author upon request.

### Application of the AGS Severity Scale in the MDBP PTPN1-related AGS cohort

The AGS Severity Scale is a disease-specific tool that measures neurologic function and correlates with the Gross Motor Function Measure-88 (GMFM-88). ^19^ The score includes evaluation for gross motor, fine motor, communication, and microcephaly; it ranges from 0 to 11. A full score of 11 is typically expected only after two years age, when subjects are expected to have achieved all the included milestones. Neurologic severity can be stratified by AGS score as severe (0–3), moderate (4–8) and mild (9–11). The AGS Severity Scale was assigned to available patient encounters by a trained child neurologist.

### Interferon (IFN) Signaling gene scores performed in the PTPN1-related AGS cohort

An IFN signature was calculated to assess the presence of autoinflammation in affected individuals. ^20,21^ Whole blood was collected in PAXgene Blood RNA tubes (PreAnalytiX) and RNA was extracted using PAXgene Blood RNA kit (Qiagen). The blood samples were obtained from the nine individuals with suspected AGS before and after the treatment of Baricitinib. Extracted RNA was quantified on Quant-iT RNA assay (Thermo) and RNA integrity number (RIN) determined using Agilent TapeStation 4200. Copy number of mRNA transcripts of previously reported type I IFN-inducible genes with increased levels in individuals with AGS and four housekeeping genes (*ALAS1*, *HPRT1*, *TBP*, and *TUBB*) was quantified using Nanostring nCounter^™^ Digital Analyzer. The raw copy number of mRNA transcripts of each type I IFN-inducible gene was standardized (stdGene) using the geometric mean of the four housekeeping genes for each individual. Then, a Z score for each IFN inducible gene was calculated using the mean and the standard deviation (SD) in the normal control group values, described in Adang et al., ^21^ according to the type of sample. In keeping with recent publications, the score for IFI44L alone (Youden’s index cut-off 5.82), a four gene score (*IFI44L*, *IFI27, USP18* and *IFI6,* Youden’s index cut-off 4.82), and the score for NIH 6 genes (*IFI27*, *IFI44*, *IFI44L*, *ISG15*, *RSAD2*, and *USP18,* Youden’s index cut-off 7.05) was assessed. The scores for IFI44L were used in primary analysis as the best score to distinguish cases as AGS from non AGS ^21^ with other scores provided in supplemental data. Amsterdam UMC cases underwent a separate ISG testing process using a 5 gene panel (*IFI44*, *IFI44L*, *IFIT1*, *IFIT3*, and *MXA*) with a Z-score cut off of 9.4 as previously described.^22^ No cases from the UCL Ion cohort underwent ISG testing.

### Exome Sequencing and Long-Read Genome Sequencing at BCM-GREGoR

BH15159-1 (LD_0474.0) and BH16506-1 (LD_2578.0) who were part of MDBP underwent research genetic evaluation as part of the BCM-GREGoR research program at the BCM Human Genome Sequencing Center (HGSC). Research trio ES was performed on BH15159-1 and his parents as previously described.^30^ BH16506-1 (LD_2578.0) and his parents underwent trio PacBio long-read genome sequencing and analysis according to a protocol developed in the BCM-HGSC (**Supplemental Methods**). ^31–34^ All genomic data generated by the BCM-GREGoR research center were deposited into the AnVIL repository in the GREGoR consortium workspace (https://anvilproject.org/). Annotated variant call files were then assessed to identify deleterious variants.

### Genome/Exome Reanalysis in the MDBP Cohort

After identification of variants in BH15159-1 (LD_0474.0) and BH16506-1 (LD_2578.0), a targeted analysis workflow was used to identify variants in *PTPN1* in the genome and exome sequence files of the unsolved AGS MDBP cohort. The aligned bam files (GRCh37) were obtained from the MDBP data repository and GATK (v4.4) HaplotypeCaller^23^ was used to produce gVCF files for each individual. A cohort-level GVCF was produced using the GATK CombineGVCF and individuals were jointly genotyped using the GATK GenotypeGVCFs protocol. The combined, cohort-level VCF was annotated using VEP 106 ^24^ for several annotations including but not limited to the functional consequence of variants, minor allele frequencies from gnomAD v3.2, ^25^ computational predictions for noncoding variants such as UTRannotator ^26^ and spliceAI, ^27^ and deleteriousness scores such as CADD ^28^ and REVEL.^29^ The annotated VCF was filtered for exonic, protein-altering variants, and intronic variants with a spliceAI > 0.2. The resulting set of variants were manually reviewed for clinical relevance.

### Additional academic cohorts

Two additional academic cohorts enriched for unsolved neurodegenerative cases were searched, the Amsterdam UMC and the UCL IoN, using data from individuals who underwent exome sequencing at local clinical or research laboratories according to established protocols.^35^

### PTPN1 variants in All of Us

To assess the prevalence, penetrance, and phenotypic spectrum associated with ultra-rare *PTPN1* pLOF and damaging missense variants in the population, we analyzed genotype–phenotype data from the *All of Us* Research Program, a large, longitudinal U.S. biobank that integrates whole-genome sequencing (WGS), electronic health record (EHR) data, survey responses, and physical measurements from a diverse adult population.^36^ We queried the Variant Annotation Table v7.1 of the *All of Us* controlled-access genomic dataset (Whole Genome Sequencing, GRCh38) for participants harboring germline *PTPN1* variants. We then annotated variants with OpenCravat (https://www.opencravat.org) and filtered down to ultra-rare predicted loss-of-function (pLOF) variants and predicted damaging missense variants using gnomAD v2.1.1 allele frequency <1/10,000 and AlphaMissense, CADD, and REVEL cutoffs ≥ ACMG/AMP PP3 Supporting thresholds (CADD≥25.3, REVEL≥0.644, AlphaMissense≥0.792).^37,38^ For each *PTPN1* variant carrier, demographic and clinical phenotypic data were extracted by joining participant identifiers to the Observational Medical Outcomes Partnership (OMOP) Common Data Model condition tables. Diagnoses were represented using standardized Systematized Nomenclature of Medicine Clinical Terms (SNOMED CT) concept identifiers mapped from source International Classification of Diseases, Tenth Revision, Clinical Modification (ICD-10-CM) codes, and all available condition records were retrieved along with their corresponding concept annotations.

### Statistical Analysis

Descriptive statistics for the cohort were calculated, including frequencies and percentages for categorical variables and means, standard deviations (SDs), medians, ranges, and interquartile ranges (IQRs) for continuous variables. Age at symptom-onset of the clinical cohort was compared to the large AGS Natural History cohort recently published ^39^ of 167 individuals with diagnosed AGS. The Mann-Whitney and ANOVA tests were used to compare the age of onset between individuals with *PTPN1* variants and those with variants in the other AGS genes. Median age of onset was also calculated for all published cases: the 13 cases in this study and the 12 previously published cases in Zhu et al. 2025.^16^ Age of onset for each subject was obtained from Table 2 in that manuscript.^16^ All statistical analyses used two-sided testing with an alpha level of 0.05. Analyses and visualizations were created using prism and R (packages/libraries loaded: survival, survminer, tidyverse, rstatix, ggplot, waffle, table1, lmtest).

## Results

### Clinical Description of PTPN1-related AGS

Within the MDBP molecularly unsolved AGS cohort, nine individuals (LD_0474/ BH15159-1, LD_1504, LD_1289, LD_1433, LD_2020, LD_2444, LD_2578/ BH16506-1, LD_2933, LD_3081) were found to have ultra-rare *PTPN1* pLOF or predicted damaging missense variants. These individuals all have biochemical markers of type 1 interferonopathy such as persistently raised CSF neopterin and ISG scores, as well as consistent clinical and radiologic features.

Neurologic and systemic manifestations were assessed across this cohort (**Table 1A and Supplemental Table 1A**). In contrast to other causes of AGS, no discernable prodrome (defined as increasing symptoms such as fevers, chilblains, irritability and myositis) was observed. ^40^ In five individuals, disease presentation followed a viral illness. Before disease onset, seven individuals were independently ambulatory and eight had at least gained single word language skills. At neurologic presentation, children had regression of speech and language, loss of motor milestones as well as development of ataxia, limb spasticity and dystonia. Disease course was notable for rapid regression with persistent neurological abnormalities.

**Table 1A:** *PTPN1* Variants and Clinical Manifestations of disease in definitive cases of *PTPN1*-related AGS.

|  |  |  |  |  |  |  |  |  |  |
| --- | --- | --- | --- | --- | --- | --- | --- | --- | --- |
|  | **LD_0474.0** | **LD_1289.0** | **LD_1433.0** | **LD_1504.0** | **LD_2020.0** | **LD_2444.0** | **LD_2578.0** | **LD_2933.0** | **LD_3081.0** |
| ***PTPN1* NM_002827.4:** | c.369del | c.595C>T | c.625G>A | c.949del | c.809_811del | c.809_811del | c.1096C>T | c.625G>A | c.456_460del |
| ***PTPN1* NM_002827.4:** | p.Gln123Hisfs<br>Ter11 | p.Arg199Ter | p.Gly209Arg | p.Leu317Trpfs<br>Ter18 | p.Ser270del | p.Ser270del | p.Gln366Ter | p.Gly209Arg | p.Tyr153Serfs<br>Ter27 |
| **MRI abnormalities** |  |  |  |  |  |  |  |  |  |
| Calcifications | + putamen | +putamen | +putamen | - | - | +putamen | +putamen | - | +putamen |
| Atrophy | +diffuse mild<br>atrophy | +diffuse<br>mild<br>atrophy | +diffuse<br>mild<br>atrophy | +diffuse mild<br>atrophy | - | +diffuse mild<br>atrophy | +diffuse mild<br>atrophy | +diffuse<br>mild<br>atrophy | +diffuse mild<br>atrophy |
| White matter signal<br>abnormalities | -delayed<br>+deep WM<br>changes | +delayed<br>+PV<br>capping | +delayed<br>+PV<br>capping | -delayed<br>+deep WM<br>changes | -delayed<br>+scattered<br>WM foci | +hypomyelin<br>ated<br>+PV capping | +delayed<br>+PV<br>capping | -delayed<br>+diffuse<br>WM<br>changes | - |
| **Biochemical testing** |  |  |  |  |  |  |  |  |  |
| Abnormal ISGs | ++ | ++ | ++ | ++ | ++ | ++ | ++ | ++ | ++ |
| CSF neopterin | 233 | >300 | 302 | 139 | 279 | >300 | 245 | 224 | 157 |
| **Neurologic features** |  |  |  |  |  |  |  |  |  |
| Acute regression | + | + | + | + | + | + | + | + | + |
| Spasticity | + | + | + | + | + | + | + | + | + |
| Axial Hypotonia | + | + | + | + | + | + | + | + | - |
| Dysphagia | + | + | + | + | + | + | + | + | + |
| Dystonia | + | + | + | + | + | + | + | + | + |
| Irritability | - | + | + | + | + | + | + | + | - |
| Ataxia | - | - | + | + | + | + | + | + | + |
| **Systemic complications** | Yes | Yes | Yes | Yes | Yes | Yes | Yes | Yes | Yes |
| **IFN blockade** | Yes | Yes | Yes | Yes | Yes | Yes | Yes | Yes | Yes |

*ND Not Done*
*+ Present*
*- Absent*

All individuals had a change in motor function, spasticity, dystonia, and dysphagia, and 8/9 had a loss of communication skills and axial hypotonia. Irritability was seen in 7/9 individuals, and ataxia and lethargy were present in 6/9 individuals. Systemic complications observed in these individuals were most commonly hematological and biochemical abnormalities. Hepatocellular injury (abnormal AST/ALT) was observed in 7/9 individuals and blood lipid abnormalities in 6/9 individuals, with two of these individuals also having abnormal Very Long Chain Fatty Acids at disease presentation. Anemia and white cell abnormalities were seen in 5/9 individuals with two additional individuals having anemia or white cell abnormalities only. The next most common systemic manifestations were inflammatory skin changes and urinary tract infections in 5/9 individuals. All systemic complications observed are detailed in Supplemental Table 1A.

All individuals had elevated CSF neopterin levels, with a median level of 233 (IQR=71.3), in some cases with recurrent abnormalities. Additionally, all individuals had at least one elevated ISG (**Figure 1B, Supplemental Table 2 and Supplemental Figures 2A&B**).

**Figure 1.**
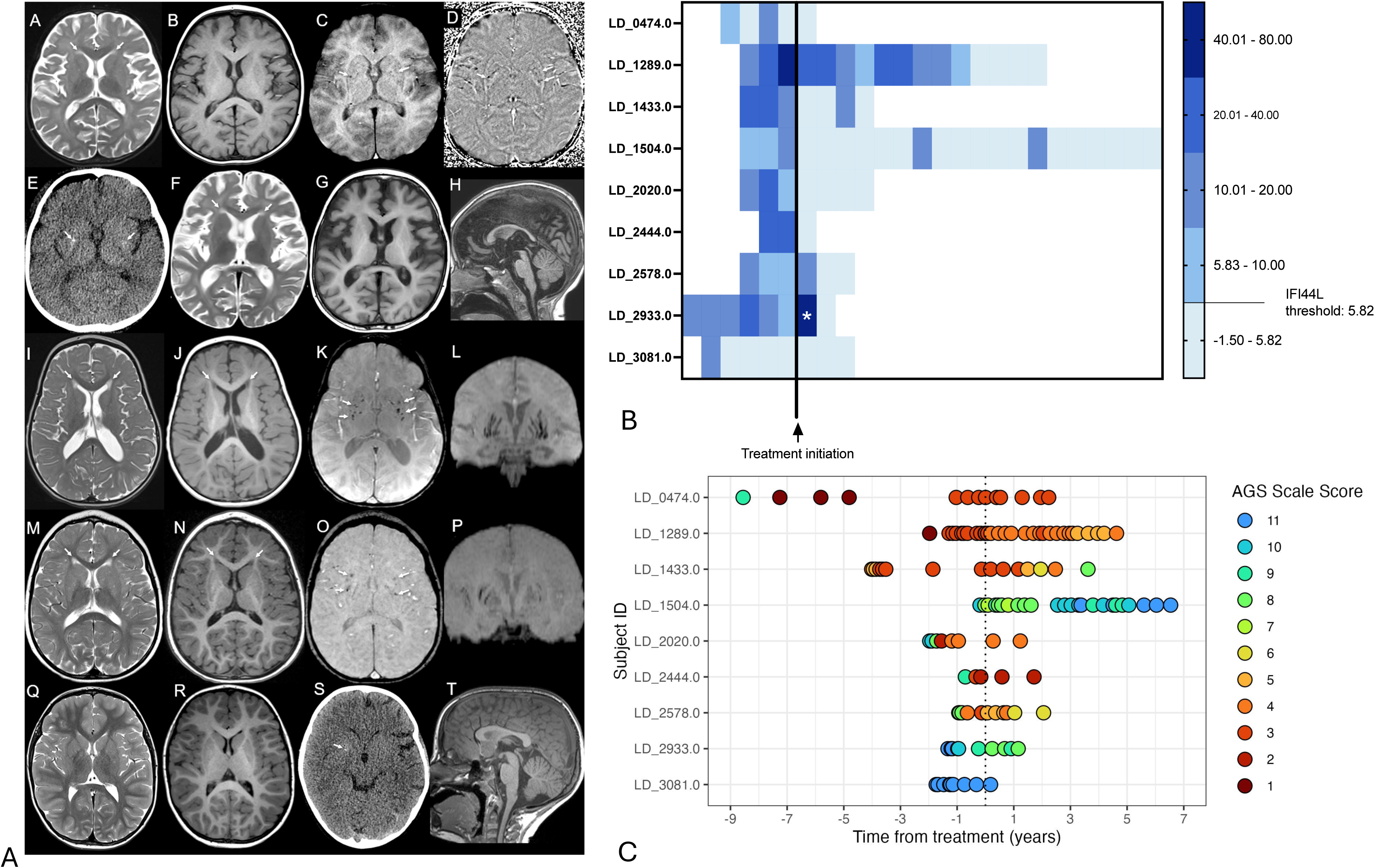
A,B&C: Imaging features, ISG scores and AGS Severity scores of patients with PTPN1-related AGS**. A** Four individuals were selected with representative imaging across different ages (LD_2444, younger in first row and older in second row, LD_1289 is in third row, LD_2578 is in fourth row and LD_3081 is in bottom row). Subjects demonstrate variable delayed or hypomyelination and volume loss (A,B with progressive changes on follow up F.G for LD_2444 or I, J for LD_1289) or in some cases normal myelination and volume (M, N for LD_2578 or Q,R for LD_3081). A subset of patients have bifrontal T2 signal abnormalities capping the frontal horns (arrows F, G, I, J, M, N) and diffuse thinning of the corpus callosum (H), but not all (Q,R, T). Susceptibility weighted imaging (C, K, O, S) and phase (D) show bilateral putamina calcifications in a subset of individuals. A CT scan performed after trauma in subject LD_2444 (E) also demonstrates the calcifications (arrows). In some individuals, coronal slab reformats (L, P) of the susceptibility imaging demonstrates that the calcifications are along the lateral lenticulostriate vessels, in keeping with a calcified lenticulostriate angiopathy. **B** IFI44L ISG scores at a particular timepoint before and after Baricitinib are represented as a color block, colored according to the scale on the right Each Score is plotted chronologically both before and after treatment initiation. All patients have at least one positive ISG score in most cases all pre treatment values are above the normal cut-off Youden value 5.82. LD_0474.0 and LD_3081.0 have some normal ISG values pre treatment. After treatment some scores fall below this normal cut off some patients scores take time to come down but all cases eventually drop below the normal cut off in all cases. The ISG score starred * for LD_2933.0 was in the context of a documented infection. **C** AGS Severity scores are depicted in a swim-lane plot where AGS severity scores at each scorable encounter are shown as a dot colored according to the scale colors shown in the key where 1 is the most severe and 11 the mildest disease. When comparing the last score prior to initiation of Baricitinib and the score at last encounter scores remained stable in LD_0474.0, LD_1289.0, LD_2020.0 and LD_2444.0 and LD_3081.0 (n=5) and improved in LD_1433.0, LD_1504.0 and LD_2578.0 and LD_2933 (n=4).

### Radiological features of PTPN1-related AGS

MRI findings are similar to those reported in prior descriptions of AGS including putaminal calcifications in 6/9, mild cerebral atrophy in 8/9, and frontal white matter hyperintensities in 8/9 **(Table 1A)**. Deep gray matter atrophy and delayed myelination were seen rarely. Detailed images for four individuals are shown in **Figure 1A** and represent the prominent features. The calcifications seen in some of these individuals appear to indicate a mineralizing angiopathy process along the lenticulostriate vessels something that has not been described previously in other AGS genotypes. Detailed MRI features in each individual are described in **Supplemental Table 3**.

### Therapeutic Response to Immunosuppressive Therapies in PTPN1-related AGS

All nine individuals were initially treated with immunosuppressive therapies, including IV methylprednisolone in five individuals and IVIG in seven individuals, based on clinical impression of an autoimmune/autoinflammatory process. At the time of this paper, all nine individuals with a clinical diagnosis of AGS are on Baricitinib (median age at treatment initiation was 3.25 years (IQR=3.35). The median time to Baricitinib treatment after disease onset in this cohort was 1.5 years (IQR=2.68). ISG scores were elevated above control level repeatedly in 8/9 prior to Baricitinib treatment. In 7/9 individuals reduction of ISG scores was observed after initiation of Baricitinib, although reduction was notably delayed in LD_1289.0 compared to other individuals. Two individuals LD_0474.and LD_3081 had scores below the cut-off at some point prior to treatment with LD_3081.0 only having one elevated score prior to initiation of IVIG and then Baricitinib (**Figure 1B**). In both cases, their progressive clinical disease determined the decision to initiate Baricitinib treatment. AGS severity scores shown in **Figure 1C** remained stable (n=5) and improved (n=4) when comparing the last score prior to initiation of Baricitinib and the score at last encounter. This response across the cohort is consistent with that observed in cases with other etiologies of AGS. ^41^ As Baricitinib was administered clinically, time intervals, the number of pre- and post-treatment ISGs and assessments of AGS severity and intervals vary between individuals; this along with the small sample size means no definitive treatment effect can be quantified.

### Family History of Autoimmunity in the PTPN1-related AGS cases

In all nine individuals with PTPN1-related AGS, the variant was inherited from an unaffected parent. To further understand this apparent lack of penetrance, a detailed family history was recorded in all individuals. As *PTPN1* and other interferonopathies have also been associated with autoimmune/autoinflammatory disorders such as Sjogren’s syndrome, systemic lupus erythematosus, Hashimoto thyroiditis, scleroderma, rheumatoid arthritis, and type 1 diabetes mellitus, the presence of these was considered a positive family history for isolated autoimmune disorders. In seven of the nine individuals, the parent from whom the variant was inherited either had an autoimmune/autoinflammatory diagnosis themselves or a first-degree relative with an immune-mediated disorder (**Supplemental Table 4**).

### Clinical Description within additional academic cohorts

In four additional individuals with *PTPN1* pLOF variants identified in the Amsterdam UMC and the UCL IoN cohorts, a definitive clinical diagnosis of AGS could not be confirmed; therefore, they were excluded from the detailed description of the *PTPN1*-related AGS but are described in **Table 1B** and **Supplemental Table 1B**.

**Table 1B:** *PTPN1* Variants and Clinical Manifestations of disease in consistent cases of PTPN1 related AGS.

|  |  |  |  |  |
| --- | --- | --- | --- | --- |
|  | **AGS-41** | **AGS-43** | **SYNS-05431** | **SYNYS-04513** |
| ***PTPN1* NM_002827.4:** | c.845delinsCG | c.505C>T | c.354+1G>T | c.645C>A |
| ***PTPN1* NM_002827.4:** | p.Met282ThrfsTer61 | p.Arg169Ter | p.? | p.Cys215Ter |
| **MRI abnormalities** |  |  |  |  |
| Calcifications | - | - | - | - |
| Atrophy | - | - | - | +diffuse mild atrophy |
| White matter signal abnormalities | -delayed<br>+confluent deep WM changes | +delayed<br>+multifocal WM changes | - | - |
| **Biochemical testing** |  |  |  |  |
| Abnormal ISGs | + | - | ND | ND |
| CSF neopterin | ND | ND | ND | ND |
| **Neurologic features** |  |  |  |  |
| Acute regression | + | + | + | + |
| Spasticity | + | + | + | + |
| Axial Hypotonia | + | + | + | - |
| Dysphagia | + | - | - | + |
| Dystonia | + | - | + | - |
| Irritability | + | - | - | + |
| Ataxia | - | - | - | + |
| **Systemic complications** | Yes | No | No | Yes |
| **IFN blockade** | No | No | No | No |

*ND – Not Done*
*+ Present*
*- Absent*

In two individuals (SYNS-04513 and SYNS-05431), no samples were available for type 1 interferon activation assays or CSF neopterin studies. AGS-41 and AGS-43 had a single ISG measurement: one elevated (AGS-41) and one normal (AGS-43). Samples from AGS-41 and AGS-43 were not available for CSF neopterin testing. However, all individuals had a clinical history consistent with neuroinflammation and reported a history of normal early development without any concerns followed by a sudden onset of neurologic features and acute developmental regression (**Tables 1B and Supplemental 1B**). A case summary of each patient is provided in Supplemental data.

### Age of onset across all subjects versus classical AGS

The median age of onset in our broader AGS natural history study is 0.33 years (IQR 0.82) with the gene having the latest age of onset being *ADAR* with median age of onset at 0.9 years (IQR 1.14). All nine *PTPN1*-related AGS patients presented with neurologic features at a median age of 1.75 years (IQR=0.67), and only one individual presented before 12 months, with the remainder presenting between 18 and 45 months of age. The age at symptom onset for this *PTPN1*-AGS cohort is significantly later than that of patients with variants in other genes known to cause AGS (1.75 years, IQR 0.67, p < 0.00001) (**Figure 2**). The median age of onset when all 13 individuals are considered is 1.97 years (IQR=1.42). The cases identified by Zhu et al 2025 had a slightly later age of onset with median of 3 years (IQR=2.88) with age of onset ranging from 15 months to 8 years of age.^16^ Aggregating the age of onset data for the 13 individuals in this paper and the 12 cases in Zhu et al. 2025 yields a median age of 2.0 years (IQR 2.32, n=25).

**Figure 2:**
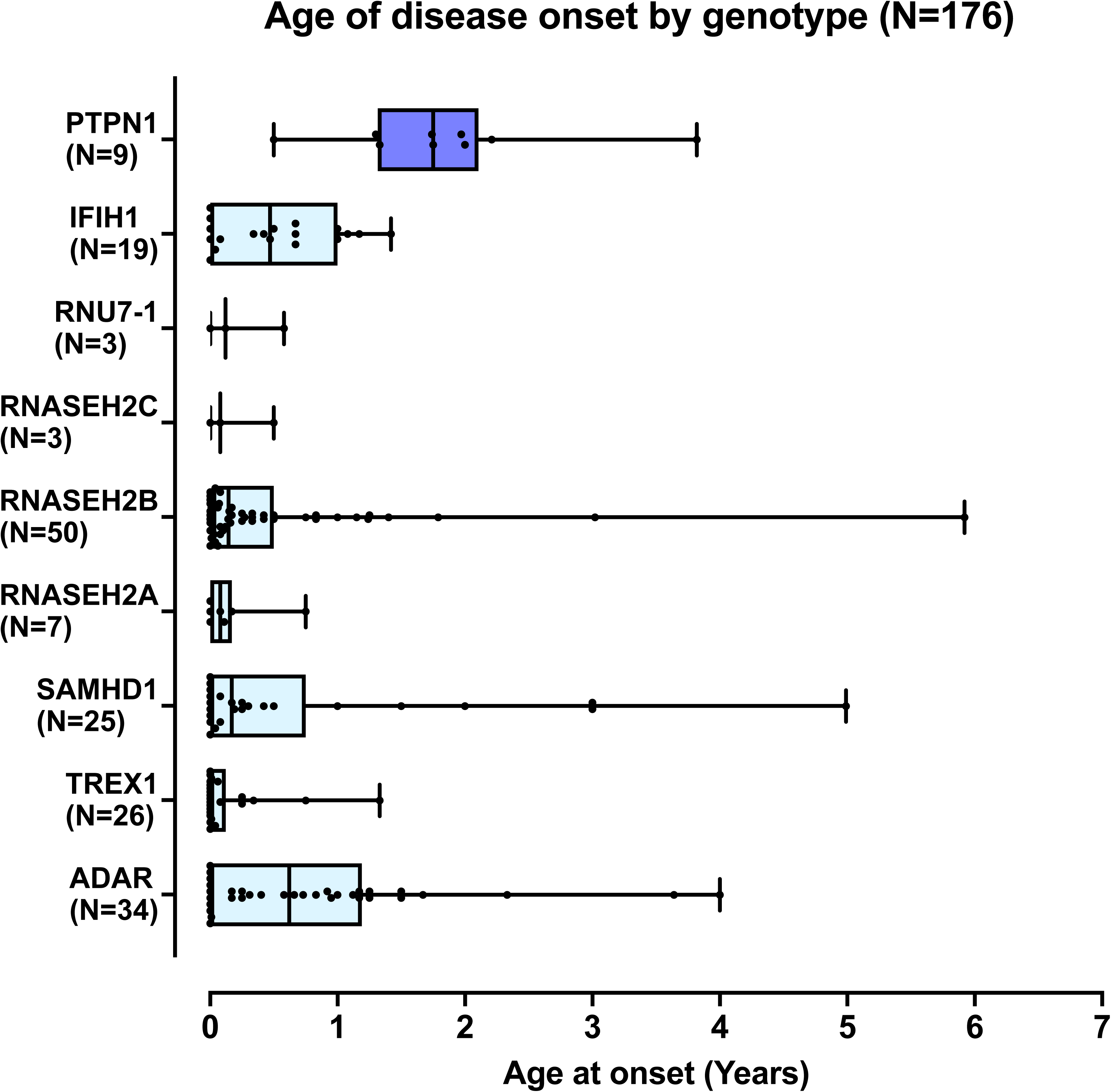
Age at symptom onset by AGS genotype. Age at symptom onset plotted for all PTPN1-related AGS cohort and confirmed AGS cases from the AGS Natural history study published^39^, divided according to their causative gene. Median age of onset for PTPN1 is significantly later (p <0.0001) than any other gene causing AGS.

### Genetic Findings

Overall, thirteen unrelated individuals, four females and nine males, were identified with private or ultra-rare *PTPN1* (NM_002827.4) pLOF or predicted damaging missense variants **(Tables 1A & B and Figure 3).** Further variant details including ACMG classifications are provided in **Supplemental Tables 5 and 6**. In two individuals, SYNS-05431 and AGS-43, the *PTPN1* variant occurred *de novo*; in all other individuals where inheritance was known, variants were inherited from an unaffected parent. In two families, grandparents were tested by Sanger sequencing (**Supplemental Figure 3**). The *PTPN1* variant c.369del, p.Gln123HisfsTer11 occurred *de novo* in the unaffected carrier parent of LD_0474.0/BH15159-1, whereas c.1096C>T, p.Gln366Ter was inherited from an unaffected carrier grandparent of LD_2578.0/BH16506-1.

**Figure 3.**
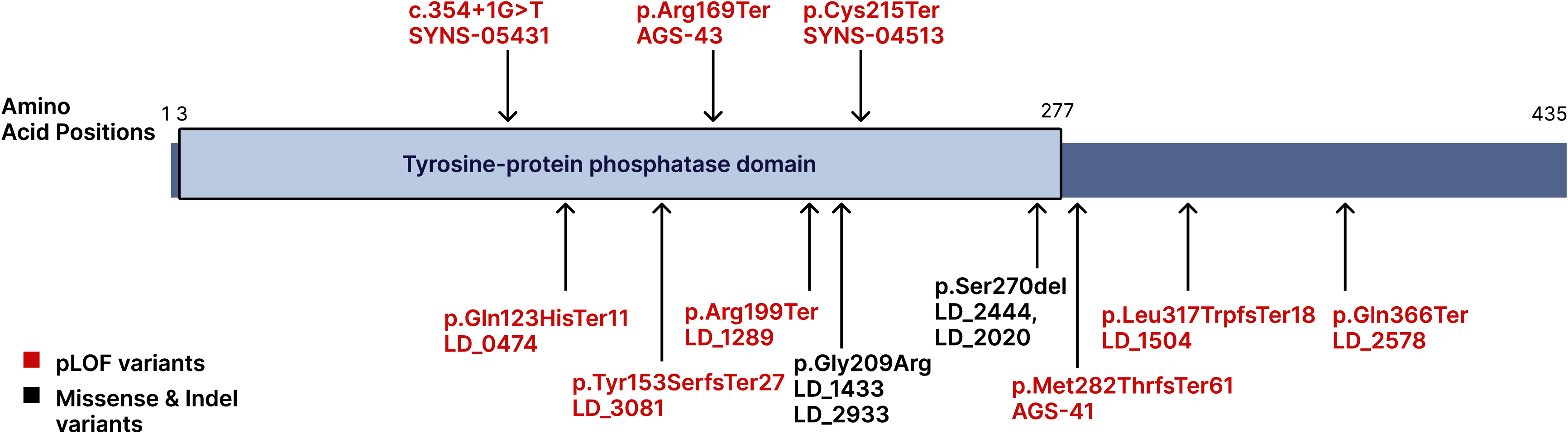
Variant location within PTP1B protein. Schematic of the PTP1B protein showing the location of the *PTPN1* variants in using HGVS p. nomenclature and HGVS c. nomenclature for the intronic splice variant. pLOF variants are colored red and missense variants black. Protein Domains taken from Uniprot **P18031.**

In total, four frameshift, four nonsense, one splice site, one in-frame deletion, and one missense variant were identified (**Figure 3**). All variants are ultra-rare or absent from gnomADv4. The in-frame indel c.809_811 del p.(Ser270del) was observed in two individuals, and the missense variant p.Gly209Arg was also observed in two individuals. The missense variant is predicted damaging by multiple *in silico* predictors; additionally, the missense variant and the in-frame deletion fall within a region with high missense constraint in gnomAD v2.1.1 (Met1-Val274, missense observed/expected 0.3255). All frameshift and nonsense variants introduce premature termination codons (PTCs) that are predicted to trigger nonsense-mediated decay (NMD) and thereby represent null alleles. *PTPN1* pLOF were significantly enriched in the undiagnosed AGS MDBP cohort (Five pLOF cases out of 53 total) versus gnomAD v2.1.1 (16 pLOF cases out of 141,456 total) (Fisher’s exact test, <0.0001). Furthermore, *PTPN1* variants potentially explain ∼17% of undiagnosed AGS cases in the MDBP cohort (9 out of 53 total).

### PTPN1 variants in All of Us

Familial segregation of *PTPN1* variants across our cohort and the published cohort ^16^ strongly supports reduced penetrance of *PTPN1*-related AGS. To further characterize the phenotypic consequences of *PTPN1* variants in the population, we analyzed *PTPN1* genotype-phenotype data in the *All of Us* database. The *All of Us* v7.1 database contains 49 participants with 31 unique ultra-rare *PTPN1* pLOF variants or damaging missense variants out of 245,394 short-read whole genome sequences. The *PTPN1* variants include 10 pLOF variants (three stopgain, seven frameshift) and 21 missense variants (**Supplemental Figure 4** and **Supplemental Table 7**). All pLOF variants and 15 missense variants are private; the most recurrent missense variant, c.793G>C p.(Asp265His), was found in seven participants. All missense variants fall within the aforementioned region of high missense constraint in gnomAD v2.1.1 (Met1-Val274). Ages of *PTPN1* variant carriers ranged from 24 to 92 years (median 50 years, standard deviation 19.1 years). Sex distribution skewed towards females (16 males, 33 females).

Phenotypic terms were available for review for 37 of the 49 participants (11 males, 26 females). None of the *All of Us* participants with *PTPN1* pLOF nor qualifying missense variants exhibited phenotypic features consistent with *PTPN1*-related AGS. As interferonopathies can exhibit variable expressivity with some individuals manifesting isolated autoimmune disorders, we also examined autoimmune phenotypes within the *All of Us PTPN1* cohort. Autoimmune phenotypic terms including Sjogren’s syndrome, lupus erythematous, Hashimoto thyroiditis, scleroderma, rheumatoid arthritis, and type 1 diabetes mellitus were identified in eight participants (∼21.6%). Recent estimates place the prevalence of autoimmune disease in the United States at 4.6% (3% of males, 6% of females),^42^ which may reflect an increased prevalence of autoimmunity in *PTPN1* carriers.

## Discussion

In this study, we identify 13 individuals with *PTPN1*-related neuroinflammatory disease from four rare disease research datasets, doubling the number of known cases with autoinflammatory brain disease attributed to this genotype. This second cohort of patients with *PTPN1*-related autoinflammation strengthens the statistical evidence for the gene-disease association of *PTPN1* with heritable type 1 interferon-driven neuroinflammation.^16^

However, nine of our cases were part of a larger cohort of molecularly unresolved AGS, suggesting that a significant percentage (17%) of AGS cases without a molecular etiology could be attributed to variants in *PTPN1.* Our cases share clinical features, biochemical levels and MRI features with prior reports of AGS phenotypes. Neurologic features seen in a majority of our cohort such as a change in motor function, spasticity, dystonia, loss of communication skills and axial hypotonia correspond to the neurological features most seen in AGS. ^39^ Similarly, systemic features such as dysphagia/feeding intolerance, hematologic abnormalities and hepatocellular injury, were prominent in patients with *PTPN1*-related AGS. All nine tested, individuals had highly elevated CSF neopterin and high ISG scores in serial ISG measurements, indicating a sustained interferonopathy present prior to treatment in most individuals.^21^ AGS MRI features such as cerebral atrophy, temporal horn dilatation and cerebral calcifications as well as cerebral white matter abnormalities with an anterior-posterior gradient ^43^ correspond to those in our cohort, albeit with milder features. The calcifications in a subset of the individuals were suggestive of a mineralizing angiopathy along the lenticulostriate vessels, a finding not previously described in other AGS genotypes. Together, these findings support the classification of clinically defined AGS.

Overall, this cohort expands the spectrum of disease associated with *PTPN1* to include AGS. A notable difference from other genetic causes of AGS is the significantly later median age of onset of *PTPN1*-related AGS compared with other AGS genotypes (**Figure 2**), similar to the cases identified by Zhu et al 2025, suggesting that *PTPN1* should be considered in late-onset AGS. Overall compared with other AGS cases, PTPN1-associated cases are milder, consistent with findings from our AGS natural history that showed a strong correlation between neurologic severity and age of onset.^39^

Importantly, the nine individuals in this case series with *PTPN1*-related AGS were treated empirically with Baricitinib based on a clinical diagnosis of AGS. JAK inhibition with Baricitinib resulted in observable reduction of ISGs in 7/9 patients. We also observe improvement of the AGS severity in some individuals and stability in others, consistent with findings in other AGS cohorts. The lack of clear natural history studies in *PTPN1*-related AGS and the small cohort size precludes determination of the effect of Baricitinib treatment in this subset of individuals. However, given the similar response seen in these individuals compared to other genotypes of AGS, we would suggest early consideration of Baricitinib treatment for patients who are identified with *PTPN1*-related AGS and persistent elevation in ISGs or other interferon biomarkers.

Our broader cohort also underscores the variability seen in *PTPN1*-related autoinflammatory disease. For example, milder disease was observed in SYNS-05431 and SYNYS-04513 whose ages of onset are 2.75 and 4, respectively. These individuals, with a later of age of onset more comparable to the Zhu *et al*. cohort, showed improvement of symptoms after their initial regression without interferon blockade similar to what was observed by Zhu *et al*. 2025. ISGs also were variable across our broader cohort, with normal ISG levels in AGS-43, LD_0474.0, and LD_3081, highlighting the importance of serial measurement of ISG scores. The frequency and significance of this finding needs further follow up and investigation in terms of what should be expected for *PTPN1* cases. Thus, prognostication remains difficult for *PTPN1*-related neuroinflammation.

Thus, a crucial point to highlight in these cases is the reduced penetrance observed in most cases; all but one variant was inherited from a parent not affected by AGS. It is challenging to estimate the penetrance of *PTPN1*-related AGS due to ascertainment bias in clinical cohorts and biobanks. Nonetheless, a range of penetrance rates can be estimated with an upper end of 50% if only clinical cohorts are considered and a lower end of ∼24% if clinical cohorts and all *All of Us* cases are aggregated (**Supplemental Table 8**). While reduced penetrance is rare but increasingly appreciated in severe neurodevelopmental disorders like *PTPN1*-related AGS ^44^, incomplete penetrance and variable expressivity have been described previously in interferonopathies ^45,,46^, immunodeficiencies ^47^ and other immune-related disorders. The variability between individuals is possibly a result of additive effects of immune gene polymorphisms ^48^ and environmental influences such as infections or may reflect compound inheritance of rare *PTPN1* loss-of-function variants and a common hypomorphic allele ^49,50^ but the small number of reported cases precludes this type of analysis. Two previous studies have shown that relatives or patients with AGS have a significantly higher rate of autoimmune diseases than controls - both studies reporting maternal predominance of this finding. ^51,52^ In this study, we also examined the frequency of other autoimmune diseases in the *All of Us PTPN1* carriers as well as personal or family history of autoimmunity in the carrier parents of our cases. We found a higher rate of autoimmunity than expected in the general population, with maternal predominance which may point to variable expressivity in some cases. Incomplete penetrance and/or variable expressivity pose significant challenges for genetic counseling in *PTPN1*-related AGS cases. Testing recommendations for unaffected siblings and relatives are unclear, and thoughtful guidance is needed to minimize the psychological impacts of this uncertainty for families.

In summary, we demonstrate that *PTPN1* pLOF variants are significantly enriched in clinically diagnosed AGS cases with no molecular etiology. Aside from a later age of onset, *PTPN1-*related AGS is almost indistinguishable from other causes of AGS, sharing clinical, biochemical, and MRI features of disease. *PTPN1* may be a significant cause of unresolved AGS and should be considered, in particular if the age of onset is after 1 year. At this stage, the efficacy of treatment with interferon blocking agents such as baricitinib remains unclear, but may be predicated by the severity of the disease presentation and earlier age at onset. *PTPN1* loss-of-function is associated with a remarkable degree of incomplete penetrance, which complicates genetic counseling and treatment, and warrants further investigation to elucidate the underlying pathomechanisms of disease.

## Author Contributions

Conceptualization: D.G.C., E.K.W., A.Va. Data Curation: E.K.W., A.S., R.D., A.P., M.D., L.E., R.M., H.H., S.N.J. Formal Analysis: D.G.C., E.K.W., F.G., L.A.A., A.S., C.D.G., M.T.W., A.Vo., R.Ri., R.Ra., M.D., Funding Acquisition: A.Va., H.H., J.R.L., K.E.S., J.E.P., R.A.G., R.Ra., D.G.C. Investigation: K.A., A.T., J.L., M.G., J.M.L., N.I.W., Q.W. Methodology: R.D., A.T., H.H.M., D.M.M., F.J.S. Project Administration: A.P., S.N.J., D.M., J.E.P., J.R.L. Resources: M.B., L.A.A., M.A., D.E.F, C.O., S.W., T.R., R.S., D.D., I.K.C, K.F., C.A.A, R.K., N.I.W Software: T.J., C.S., R.Ri., E.F., R.Ra. Supervision: A.Va., L.A.A., K.E.S., H.H., J.E.P., J.R.L, R.A.G. Visualization: F.G., S.W. Writing original draft: D.G.C., E.K.W., M.D. Writing Review & editing: D.G.C., E.K.W., F.G., A.S., A.P., K.A., C.D.G., T.J., M.B., L.A., S.W., M.T.W., A.Vo., R.D., A.T., J.L., C.S., R.Ri., E.F., K.E.S., M.A., D.E-F., C.O., S.W., T.R., N.I.W., Q.W., D.D., M.D., M.G., J.M.L., I.K.C., K.F., L.E., C.A.A., R.K., R.M., H.H., S.N.J., H.H.M., D.M.M., F.J.S., J.E.P., J.R.L., R.A.G., R.Ra. A.Va

## Ethics Declaration

Ethics approval was provided by BCM IRB H-29697, CHOP IRB #14–011236, AUMC IRB #2018.300 and UCL IoN IRB #07/Q0512/26.

## Data Availability

All data produced in the present study are available upon reasonable request to the authors. All genomic data generated by the BCM-GREGoR research center were deposited into the AnVIL repository in the GREGoR consortium workspace (https://anvilproject.org/).

