## Supplemental Methods, Figures, and Tables for "*PTPN1*-related autoinflammation is a common cause of Aicardi-Goutières Syndrome with reduced penetrance"

**Supplemental Data**

1. **Supplemental Methods**
   a. BCM Gregor Long Read Sequencing Refined Methodology
2. **Supplemental Figures**
   a. Supplemental Figure 1: The role of PTPN1 in the intracellular Interferon signaling pathway
   b. Supplemental Figure 2: PTPN1-related AGS patient 4 gene ISG scores
   c. Supplemental Figure 3: Sanger Results for LD_0474.0/BH15159-1 and LD_2578.0/BH16506-1
   d. Supplemental Figure 4: Ultra-rare, predicted damaging *PTPN1* variants identified in the All of Us Database
3. **Supplemental Tables**
   a. Supplemental Table 1A&B: Systemic manifestations of disease in patients with definitive biochemically confirmed AGS(A) and without biochemical confirmation of AGS (B)
   b. Supplemental Table 2: Raw z-score values for all ISG scores for IFI44L, 4 gene, NIH6 gene AGS 6 gene - See excel document
   c. Supplemental Table 3: Detailed MRI findings for each clinical case of PTPN1-related AGS
   d. Supplemental Table 4: Details of family history in nine cases of PTPN1-related AGS cases
   e. Supplemental Table 5: *PTPN1* (*NM_002827.4)* variants identified in case with clinically consistent with AGS
   f. Supplemental Table 6: Details of ACMG criteria applied to each variant in *PTPN1* (NM_002827.4)
   g. Supplemental Table 7: *PTPN1* pLOF or damaging missense variants identified in the All of Us database and Phenotypes for each corresponding individual - See excel document
   h. Supplementary Table 8: Calculations of *PTPN1* Penetrance
4. **Clinical Vignettes**
1. **Supplemental Methods**

*BCM Gregor Long Read Sequencing Refined Methodology*

PacBio long-read genome sequencing was performed at BCM GREGOR according to the below modified protocol. Briefly, DNA quality was assessed using Qubit dsDNA quantification broad range assay (Thermo Fisher Scientific) and Agilent Femto Pulse. One library per sample was constructed starting with 7.5 µg input genomic DNA. Covaris g-tubes (Covaris 520079) were used for shearing DNA. Following bead purification using 1X Pacbio SMRTbell cleanup beads, the size of the sheared DNA was determined on the Femto Pulse. Small fragments were eliminated using size-selection on the PippinHT instrument (Sage Science) using either the 6-10 kb or the 15-20 kb High-Pass definition where the minimum size selection threshold varied based on the average size of the sheared sample. Loading and elution of samples from the PippinHT cassette were performed following the manufacturer’s instructions. The selected size samples were cleaned up using 1X Pacbio SMRTbell Cleanup Beads and eluted in 47 µL of Pacbio Elution Buffer. This was used as input into the Pacbio SMRTbell® prep kit 3.0 (PN: 102-182-700) and final libraries were constructed following the manufacturer’s instructions. Libraries were barcoded using the SMRTbell barcoded adapter index plate 96A (PN 102-009-200). Final libraries were quantified using the Qubit dsDNA HS (High Sensitivity) Assay Kit and their size was determined by the Agilent Femto Pulse. Using the Pacbio Revio instrument, one SMRT Cell was loaded per library achieving an average of 35.9x coverage and 15.8Kb per sample. Using PRINCESS version 2.0, a workflow for long read sequence analysis, reads were aligned to GRCh38 and phased variant calls were generated for SVs and SNVs.^1^ PRINCESS aligns reads using the appropriate parameters based on the type of sequencing technology using Minimap2 version 2.24 followed by calling SVs using Sniffles version 2.0.5 and identify SNVs and indels using Clair3 version 0.1.11.^2-4^  Finally, PacBio HiFi data was processed using the same methods using PRINCESS with the read-option set to CCS (--ReadType ccs). For SVs from both sequencing platforms, variants were filtered based on read support to require a maximum ∼25k SVs per sample.

All genomic data generated by the BCM-GREGoR research center were deposited into the AnVIL repository in the GREGoR consortium workspace (https://anvilproject.org/).

**Supplemental Figures**

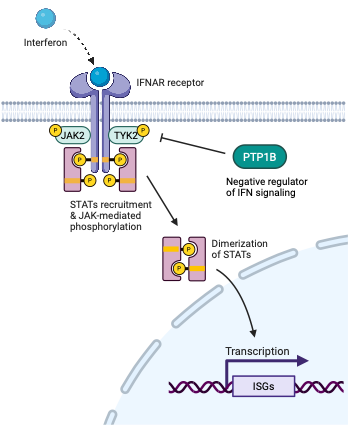

**Supplemental Figure 1: The role of PTPN1 in the intracellular Interferon signaling pathway** Schematic depicting the Interferon signaling pathway showing the location where PTP1B (the protein encoded by *PTPN1*) exerts its effect; by negatively regulating phosphorylation of JAK2 and TYK2. Loss of negative regulation by PTP1B from damaging variants in *PTPN1* results in uncontrolled IFN signaling and increased transcription of interferon signaling genes. Created in BioRender. Wiener, E. (2026) <https://BioRender.com/cwaski6>

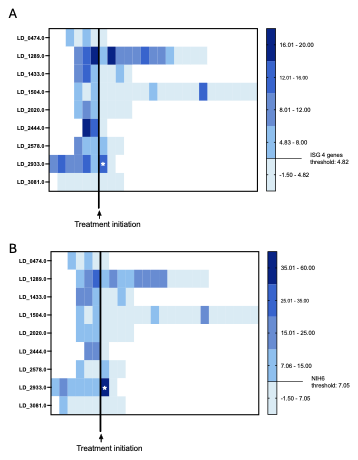

**Supplemental Figure 2A&B: PTPN1-related AGS patient 4-gene and NIH6-gene ISG scores.** Four gene ISG scores in **A** and NIH six gene scores in **B** at a particular timepoint before and after Baricitinib are represented as a color block, colored according to the scale on the right Each Score is plotted chronologically both before and after treatment initiation. four gene score (*IFI44L*, *IFI27, USP18* and *IFI6,* Youden’s index cut-off 4.82*),* and the score for NIH 6 genes (*IFI27*, *IFI44*, *IFI44L*, *ISG15*, *RSAD2*, and *USP18,* Youden’s index cut-off 7.05)

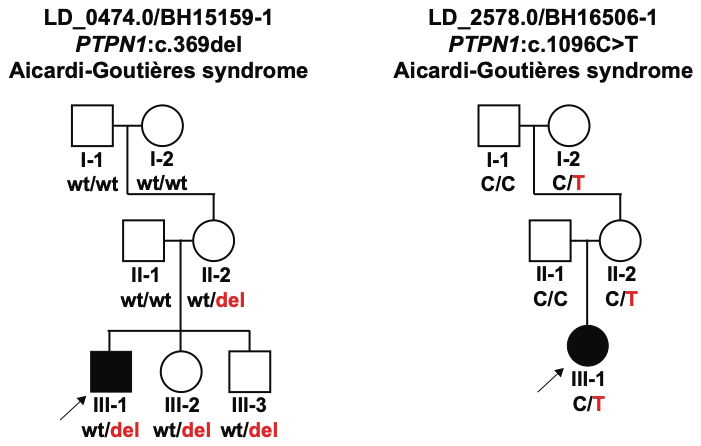
**Supplemental Figure 3: Sanger Results for LD_0474.0/BH15159-1 and LD_2578.0/BH16506-1**

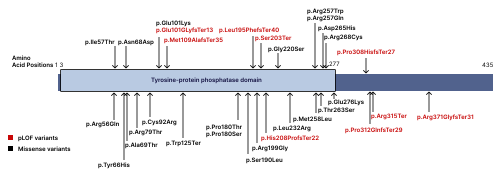

**Supplemental Figure 4: Ultra-rare, predicted damaging *PTPN1* variants identified in the All of Us Database.** Schematic of the *PTP1B* protein showing the location of the variant according to amino acid position HGVS p. nomenclature. pLOF variants are colored red and missense variants black. Domains taken from Uniprot **P18031.**

| **Supplemental Table 1A: Clincial manifestations of disease in patients with definitive biochemically confirmed AGS** | | | | | | | | | | |
| --- | --- | --- | --- | --- | --- | --- | --- | --- | --- | --- |
|  |  | *LD_0*474*.0* | *LD_*1289*.0* | *LD_*1433*.0* | *LD_*1504*.0* | *LD_*2020*.0* | *LD_*2444*.0* | *LD_*2578*.0* | *LD_*2933*.0* | *LD_*3081*.0* |
| System | Clinical Event |  | | | | | | | | |
| Endocrine Complications | Hypothyroidism | *+* |  |  | *+* |  |  |  |  |  |
|  | Diabetes Insipidus |  |  |  |  |  | *+* |  |  |  |
| Genitourinary (GU) Complications | Incontinence or Loss of Urinary Function | *+* | *+* |  | *+* |  |  |  | *+* |  |
|  | Urinary Tract Infection |  | *+* | *+* | *+* |  |  | *+* | *+* |  |
|  | Renal Complications |  |  | *+* |  |  |  |  |  |  |
|  | Neurogenic Bladder |  |  |  |  |  |  | *+* |  |  |
|  | Prophylactic Antibiotics for Chronic Kidney or Bladder Disease |  |  |  |  |  | *+* |  |  |  |
| Gastrointestinal (GI) Complications | Liver Dysfunction or Hepatocellular Injury | *+* |  | *+* | *+* | *+* | *+* | *+* | *+* |  |
|  | Dysphagia or Feeding Intolerance | *+* | *+* | *+* | *+* | *+* | *+* | *+* | *+* | *+* |
|  | Feeding Tube Placement | *+* |  | *+* |  |  | *+* |  |  | *+* |
|  | Complete Dependence on Gastric Feeds | *+* |  | *+* |  |  |  |  |  |  |
| Hematologic and Immune Complications | Autoantibody Production | *+* |  |  |  |  | *+* |  |  |  |
|  | Anemia | *+* | *+* |  | *+* | *+* |  |  | *+* |  |
|  | White Blood cell Abnormalities | *+* | *+* | *+* | *+* |  |  |  | *+* |  |
| Miscellaneous (AGS-specific) | Blood Lipid Abnormalities |  | *+* |  | *+* | *+* | *+* | *+* | *+* |  |
|  | Sterile Pyrexia | *+* |  | *+* |  |  |  | *+* |  |  |
| Musculoskeletal Complications | Myopathy, Myositis, and/or Myalgia |  | *+* |  |  |  | *+* |  | *+* |  |
|  | Arthropathy |  |  |  |  |  |  |  | *+* |  |
| Neurologic Function and Complications | Head Circumference Abnormalities |  | *+* |  |  |  |  | *+* |  |  |
|  | Spasticity | *+* | *+* | *+* | *+* | *+* | *+* | *+* | *+* | *+* |
|  | Axial Hypotonia | *+* | *+* | *+* | *+* | *+* | *+* | *+* | *+* |  |
|  | Dystonia | *+* | *+* | *+* | *+* | *+* | *+* | *+* | *+* | *+* |
|  | Irritability |  | *+* | *+* | *+* | *+* | *+* | *+* | *+* |  |
|  | Early Gross Motor Delay |  | *+* |  |  |  |  |  |  |  |
|  | Early Cognitive Delay | *+* | *+* |  |  |  |  |  |  | *+* |
|  | Change in Motor Function | *+* | *+* | *+* | *+* | *+* | *+* | *+* | *+* | *+* |
|  | Lethargy |  |  | *+* | *+* | *+* |  | *+* | *+* |  |
|  | Loss of Communication Function | *+* |  | *+* | *+* | *+* | *+* | *+* | *+* | *+* |
|  | Ataxia |  |  | *+* | *+* | *+* | *+* | *+* | *+* | *+* |
| Respiratory and Cardiac Complications | Hypertension | *+* |  |  |  |  | *+* | *+* |  |  |
|  | Obstructive Sleep Apnea (OSA) | *+* |  | *+* |  | *+* |  |  |  |  |
|  | Cardiovascular Issues |  | *+* | *+* |  | *+* |  | *+* |  |  |
| Seizure Complications | Seizure | *+* |  | *+* |  |  |  | *+* |  |  |
|  | Anti-Epileptic Medication | *+* |  | *+* |  |  |  |  |  |  |
| Skin Complications | Inflammatory Skin Changes |  | *+* | *+* | *+* |  |  | *+* | *+* |  |
|  | Chilblains |  |  |  |  | *+* | *+* |  |  |  |
| Visual Complications | Retinopathy |  |  |  |  | *+* |  |  |  |  |

**Supplemental Table 1B: Clinical manifestation of disease in cases without biochemical confirmation of AGS**

| System | Clinical Event | AGS-41 | AGS-43 | *SYNS-05431* | *SYNYS-04513* |
| --- | --- | --- | --- | --- | --- |
| Endocrine Complications | Insulin Dependent Diabetes Mellitus |  |  |  |  |
|  | Hypothyroidism |  |  |  |  |
|  | Diabetes Insipidus |  |  |  |  |
| Genitourinary (GU) Complications | Incontinence or Loss of Urinary Function | *+* |  |  | *+* |
|  | Urinary Tract Infection |  |  |  | *+* |
|  | Renal Complications |  |  |  |  |
|  | Neurogenic Bladder |  |  |  |  |
|  | Prophylactic Antibiotics for Chronic Kidney or Bladder Disease |  |  |  |  |
| Gastrointestinal (GI) Complications | Liver Dysfunction |  |  |  |  |
|  | Dysphagia or Feeding Intolerance | *+* |  |  | *+* |
|  | Feeding Tube Placement |  |  |  |  |
|  | Complete Dependence on Gastric Feeds |  |  |  |  |
| Hematologic and Immune Complications | Autoantibody Production |  |  |  |  |
|  | Anemia |  |  |  |  |
|  | White Blood cell Abnormalities |  |  |  |  |
| Miscellaneous (AGS-specific) | Blood Lipid Abnormalities |  |  |  |  |
|  | Sterile Pyrexia |  |  |  | *+* |
| Musculoskeletal Complications | Myopathy, Myositis, and/or Myalgia |  |  |  | *+* |
|  | Arthropathy |  |  |  |  |
| Neurologic Function and Complications | Head Circumference Abnormalities |  |  |  |  |
|  | Axial Hypotonia | *+* | *+* | *+* |  |
|  | Spasticity | *+* | *+* | *+* | *+* |
|  | Dystonia |  |  | *+* |  |
|  | Irritability | *+* |  |  | *+* |
|  | Early Gross Motor Delay |  |  |  |  |
|  | Early Cognitive Delay |  |  |  |  |
|  | Change in Motor Function | *+* | *+* | *+* | *+* |
|  | Lethargy |  |  |  |  |
|  | Loss of Communication Function | *+* |  | *+* | *+* |
|  | Ataxia |  |  |  | *+* |
|  | Peripheral Neuropathy |  |  |  |  |
| Respiratory and Cardiac Complications | Hypertension |  |  | *+* |  |
|  | Obstructive Sleep Apnea (OSA) |  |  |  |  |
|  | Cardiovascular Issues |  |  |  |  |
| Seizure Complications | Seizure |  |  |  |  |
|  | Anti-Epileptic Medication |  |  |  |  |
| Skin Complications | Inflammatory Skin Changes | *+* |  |  |  |
|  | Chilblains |  |  |  |  |
| Visual System Complications | Retinopathy |  |  |  |  |

**Supplemental Table 2: Raw z-score values for all ISG scores for IFI44L, four gene, NIH six gene AGS six gene**

See Excel document Sheet 1

**Supplemental Table 3: Detailed MRI findings for each clinical case of PTPN1-related AGS**

| LD | **Calcifications** | **Atrophy** | **Deep Gray Atrophy** | **WM Hyperintensity** | **Delayed Myelination** |
| --- | --- | --- | --- | --- | --- |
| LD_0474.0 | Bilateral putaminal calcifications | Initially mild cerebral atrophy only, progressed to moderate cerebral and mild brainstem/cerebellum | Yes | Foci in bilateral frontal and parietotemporal periatrial predominant; asymmetric in the frontal regions. L>R | No |
| LD_1289.0 | Putaminal calcifications vs prominent vessels (MRI only) | Initially mild cerebral only; progressed to mild cerebral and brainstem volume loss. | No | Bifrontal caps | Yes |
| LD_1433.0 | Putaminal calcifications vs prominent vessels (MRI only) | Initially mild cerebral only, progressed to mild cerebral and brainstem volume loss | No | Bifrontal caps | Yes |
| LD_1504.0 | No | Mild cerebral atrophy only | No | Faint, frontal> parietotemporal periatrial/subcortical foci; mild asymmetry in frontal regions R>L | No |
| LD_2020.0 | No | None | No | Scattered WM foci that went away, but bifrontal caps at frontal horns remained | No |
| LD_2444.0 | Bilateral putaminal calcifications | Mild volume loss over a short period | No | Diffuse hypomyelination, with bifrontal caps superimposed | Yes |
| LD_2578.0 | Putaminal calcifications vs prominent vessels (MRI only) | No atrophy initially, lack of appropriate growth by results in mild cerebral hypoplasia with mild thinning of corpus callosum. | No | Bifrontal caps | Mild |
| LD_2933.0 | No | Mild volume loss over time | No | Widespread confluent supratentorial WM T2 and T1 signal abnormality, improved (mostly) over time | No (the initial WM change is not delayed myelination) |
| LD_3081.0 | Right putaminal calcifications | Mild left-sided atrophy, best see on initial CT, not present on subsequent MRIs | No | Normal | No |

**Supplemental Table 4: Details of family history in nine cases of PTPN1-related AGS**

| Case ID | Family History | Considered as positive for autoimmunity |
| --- | --- | --- |
| LD_0474.0 | Heterozygous mother and their uncle have vitiligo | Yes |
| LD_1289.0 | Heterozygous father has eczema and irritable bowel syndrome | No |
| LD_1433.0 | Heterozygous mother has history of psoriasis and hypothyroidism | Yes |
| LD_1504.0 | Heterozygous mother has multiple sclerosis | Yes |
| LD_2020.0 | Heterozygous father - Nothing to note | No |
| LD_2444.0 | Heterozygous father and their mother have systemic lupus erythematosus | Yes |
| LD_2578.0 | Heterozygous mother has hypothyroidism | Yes |
| LD_2933.0 | Heterozygous father’s sister has type 1 diabetes | Yes |
| LD_3081.0 | Heterozygous mother's mother had severe rheumatoid arthritis | Yes |

**Supplemental Table 5: *PTPN1*** ([***NM_002827.4***](https://www.ncbi.nlm.nih.gov/nuccore/NM_002827.4)***)* variants identified in cases clinically consistent with AGS**

| Case ID | Sex | HGVS c. | HGVS p. | Inheritance | Consequence | gnomAD AF | CADD | REVEL | Alpha  missense | SpliceAI | ACMG^a^  Classification |
| --- | --- | --- | --- | --- | --- | --- | --- | --- | --- | --- | --- |
| AGS-41 | Male | c.845delinsCG | p.Met282ThrfsTer61 | Unknown | Stop gain | Absent | NA | NA | NA | NA | Pathogenic |
| AGS-43 | Male | c.505C>T | p.Arg169Ter | De novo | Stop gain | 0.00006% | 38 |  |  |  | Pathogenic |
| LD_0474.0 | Male | c.369del | p.Gln123HisfsTer11 | Maternal | Frameshift | Absent | NA | NA | NA | NA | Pathogenic |
| LD_1289.0 | Male | c.595C>T | p.Arg199Ter | Paternal | Stop gain | 0.0002% | 47 | NA | NA | NA | Pathogenic |
| LD_1433.0 | Male | c.625G>A | p.Gly209Arg | Maternal | Missense | 0.0002% | 27.6 | 0.9 | 0.874 | NA | Likely Pathogenic |
| LD_1504.0 | Female | c.949del | p.Leu317TrpfsTer18 | Maternal | Frameshift | Absent | NA | NA | NA | NA | Pathogenic |
| LD_2020.0 | Male | c.809_811del | p.Ser270del | Paternal | In-frame del | Absent | NA | NA | NA | NA | Likely Pathogenic |
| LD_2444.0 | Male | c.809_811del | p.Ser270del | Paternal | In-frame del | Absent | NA | NA | NA | NA | Likely Pathogenic |
| LD_2578.0 | Male | c.1096C>T | p.Gln366Ter | Maternal | Stop gain | Absent | 46 | NA | NA | NA | Pathogenic |
| LD_2933.0 | Female | c.625G>A | p.Gly209Arg | Paternal | Missense | 0.0002% | 27.6 | 0.9 | 0.874 | NA | Likely Pathogenic |
| LD_3081.0 | Female | c.456_460del | p.Tyr153SerfsTer27 | Maternal | Frameshift | Absent | NA | NA | NA | NA | Pathogenic |
| SYNS-05431 | Male | c.354+1G>T | p.? | De novo | Splice | Absent | 36 | NA | NA | 1 | Pathogenic |
| SYNS-04513 | Female | c.645C>A | p.Cys215Ter | Paternal | Stop gain | Absent | 37 | NA | NA | NA | Pathogenic |

a) ACMG Criteria applied detailed in Supplementary Table 2

**Supplemental Table 6: Details of ACMG criteria applied to each variant in *PTPN1* (NM_002827.4)**

| Case_ID | HGVS c. | HGVS p. | PVS1- very strong | PS2 - strong | PS4 -moderate | PM1 -moderate | PM2 – supporting^a^ | PM4 -moderate | PP3-supporting | PP4 -moderate^b^ | Classification |
| --- | --- | --- | --- | --- | --- | --- | --- | --- | --- | --- | --- |
| AGS-41 | c.845delinsCG | p.Met282ThrfsTer61 | Met |  |  |  | Met |  |  |  | Pathogenic |
| AGS-43 | c.505C>T | p.Arg169Ter | Met | Met |  |  | Met |  |  |  | Pathogenic |
| LD_0474.0 | c.369del | p.Gln123HisfsTer11 | Met |  |  |  | Met |  |  | Met | Pathogenic |
| LD_1289.0 | c.595C>T | p.Arg199Ter | Met |  |  | Met | Met |  |  | Met | Pathogenic |
| LD_1433.0 | c.625G>A | p.Gly209Arg |  |  | Met | Met | Met |  | Met | Met | Likely Pathogenic |
| LD_1504.0 | c.949del | p.Leu317TrpfsTer18 | Met |  |  |  | Met |  |  | Met | Pathogenic |
| LD_2020.0 | c.809_811del | p.Ser270del |  |  | Met | Met | Met | Met |  | Met | Likely Pathogenic |
| LD_2578.0 | c.1096C>T | p.Gln366Ter | Met |  |  |  | Met |  |  | Met | Pathogenic |
| LD_3081.0 | c.456_460del | p.Tyr153SerfsTer27 | Met |  |  |  | Met |  |  | Met | Pathogenic |
| LD_2933.0 | c.625G>A | p.Gly209Arg |  |  | Met | Met | Met |  | Met | Met | Likely Pathogenic |
| LD_2444.0 | c.809_811del | p.Ser270del |  |  | Met | Met | Met | Met |  | Met | Likely Pathogenic |
| SYNS-05431 | c.354+1G>T |  | Met | Met |  |  | Met |  |  |  | Pathogenic |
| SYNS-04513 | c.645C>A | p.Cys215Ter | Met |  |  |  | Met |  |  |  | Pathogenic |

a) PM2 applied for variants with AF <0.001%

b) PP4 applied for persistently elevated CSF Neopterin and ISGs considered specific for AGS

**Supplemental Table 7: *PTPN1* pLOF or damaging missense variants identified in the All of Us database and Phenotypes for each corresponding individual**

See excel document Sheet 2

**Supplemental Table 8: Calculations of *PTPN1* Penetrance**

| **Clinical Cohort Cases + All AoU cases with qualifying variants** | | |
| --- | --- | --- |
| *PTPN1* variant carriers | Affected | Unaffected |
| This manuscript | 12 | 14 |
| Zhu et al 2025 | 11 | 9 |
| All of Us (All Qualifying Variants) | 0 | 49 |
| Total | 23 | 72 |
| Penetrance | 0.24210526 |  |
| **Clinical Cohort Cases + All AoU cases with qualifying variants and phenotypic terms available** | | |
| *PTPN1* variant carriers | Affected | Unaffected |
| This manuscript | 12 | 14 |
| Zhu et al 2025 | 11 | 9 |
| All of Us (All Qualifying Variants + Phenotypic Terms Available) | 0 | 37 |
| Total | 23 | 60 |
| Penetrance | 0.27710843 |  |
| **Clinical Cohort Cases + All AoU cases with pLOF variants** | | |
| *PTPN1* variant carriers | Affected | Unaffected |
| This manuscript | 12 | 14 |
| Zhu et al 2025 | 11 | 9 |
| All of Us (pLOF only) | 0 | 10 |
| Total | 23 | 33 |
| Penetrance | 0.41071429 |  |
| **Clinical Cohort Cases + All AoU cases with pLOF variants and phenotypic terms available** | | |
| *PTPN1* variant carriers | Affected | Unaffected |
| This manuscript | 12 | 14 |
| Zhu et al 2025 | 11 | 9 |
| All of Us (pLOF only) | 0 | 7 |
| Total | 23 | 30 |
| Penetrance | 0.43396226 |  |
| **Clinical Cohort Cases only** | |  |
| *PTPN1* variant carriers | Affected | Unaffected |
| This manuscript | 12 | 14 |
| Zhu et al 2025 | 11 | 9 |
| Total | 23 | 23 |
| Penetrance | 0.5 |  |

3 **Clinical PTPN1-related AGS case Vignettes**

Due to Medrxiv requirements, this section has been omitted. Readers may contact the corresponding author for access to this data.
